# Characterization of the gene expression profile in younger and older low-grade glioma patients

**DOI:** 10.1101/2022.02.09.22270731

**Authors:** Cathryn Dong, Xiang Fei, Youtang Guo, Minde Jiang, Yifan Mao, Zhaoying Wang, Yutong Wei, Tianyi Yu, Yong Shi, Yi Yang, Tao Qing, Sibo Zhu

## Abstract

Low-grade gliomas (LGG) are the slowest growing type of brain cancer that often affects young adults. Of all LGG cases, older patients were associated with the poor prognosis compared to younger patients. However, the molecular mechanism underlying this association remains unclear. Here, we compared genes expression profiles between younger (age≤50) and older (age > 50) patients from The Cancer Genome Atlas (TCGA) and demonstrate the age-related gene expressions. Pathway and gene set enrichment analysis reveal that differentially expressed genes (DEGs) between tumors of older and younger patients have been involved in cancer-related biological procedures, such as, transcriptional misregulation in cancer, nucleotide synthesis, mTOR, and DNA damage repair signaling. We also demonstrated that older patients have higher expression of Mitosis Kinase Score (MKS) and Tumor Inflammation Signature (TIS) which reflects high cell proliferation and high immune cell activity; respectively. The comprehensive characterization of gene-expression profiles in younger versus older LGG may explain the age-related prognosis and facilitate the development of potential therapeutic biomarkers for LGG.

## Introduction

LGGs are a diverse group of primary brain tumors that often arise in young. LGGs generally have an indolent course with longer-term survival in comparison with high-grade gliomas. Despite a similar histologic appearance to adult LGG, the risk of higher-grade glioma increases by age [1]. Younger LGGs have a more favorable clinical outcome, with a notably lower incidence of malignant transformation. Age also affects the treatment decision of LGG [2]. More neurological deficits were observed in older patients after surgery [3] and treatment recommendations should take into account the patients’ age [4]. However, the molecular mechanism under this age-related difference remains unknown.

The genomic and transcriptomic analysis allows characterizing the molecular alterations related to the development and progression of LGG. IDH mutations were identified in >80% of tumors histologically classified as LGG [5, 6]. The larger of these subclasses is characterized by inactivating mutations in the tumor suppressor TP53 and the chromatin remodeling enzyme ATRX [7]. EN1 and CHI3L1 expression were associated with LGG prognosis which predicts a worse survival (9-10). However, it remains unclear whether those molecular changes are related to the patient’s age.

Here, we integrated RNA expression profiles of 504 patients from The Cancer Genome Atlas (TCGA), and 350 of them are younger patients (age<50), and 154 older patients (age>50). We performed differentially expression analysis by compare gene expressions of younger versus older patients, further mapped those differentially expressed genes to the Kyoto Encyclopedia of Genes and Genomes (KEGG) pathways. In order to investigate the role of metabolic genes and tumor immune microenvironment in LGGs, we compared the gene expression of 36 NanoString metabolic pathways and 6 immune signatures between younger and older patients. Older patients with LGG have higher metabolic and immune activity than that in younger patients. Our study characterizes the age-related gene-expression alterations in LGGs suggesting the mechanism of better survival in younger LGGs.

## Methods

### RNA-seq dataset

RNA-seq expression data and corresponding clinical data of 504 primary low-grade glioma patients were obtained from The Cancer Genome Atlas (https://gdc.cancer.gov/about-data/publications/pancanatlas) [8]. The expression matrix of Fragments per Kilobase of transcript per Million mapped reads (FPKM) was normalized by upper quartile which the FPKM value was divided by the 75th percentile value for the samples. Then, the matrix was transformed to log2-scale.

### Gene-expression signatures

Gene expression signatures were obtained from literature. For each signature, we calculated the average expression of the member genes with the normalized RNA-seq data and transformed to z-score across all cases. We compared the expression of each signature between young versus older cases. Fold change and Mann-Whitney test p-value were calculated. The false discovery rate (FDR) for multiple comparison adjustment was calculated using the Benjamini Hochberg algorithm.

### Differentially expressed genes

To identify differentially expressed genes, we calculated fold change and t-test p-value for each gene between younger and older cases. Differentially expressed genes were defined as fold change ≥ 1.50 or ≤ 0.67 with Benjamini Hochberg corrected false discovery rate (FDR)□<□0.05. In this analysis, we included 20,282 human genes which are available in the TCGA RNA-seq dataset.

### Gene set enrichment analysis

The Kyoto Encyclopedia of Genes and Genomes (KEGG) pathway was used to perform enrichment analysis. P value was generated by Fisher’s exact test using the “clusterProfiler” package in R.

### Gene-expression signatures

We constructed six gene-signatures (Supplementary Table1) including one proliferation signature (Mitosis Kinase Score, MKS) [9] five immune cell signatures (e.g. T Cell, B Cell, Mast Cell, Dendritic Cell [10], and Tumor inflammation signature (TIS) [11]. Metabolic genes were obtained from NanoString Technology. For each signature, we calculated the average normalized expression of the member genes and transformed to z-score across all cases in a given cohort.

### Statistical analysis

Fold change and Student’s t-test was used for differential expression analysis. Mann-Whitney test was used to compare the expression of signatures. All statistical tests were two-sided and FDR for multiple comparisons was calculated using the Benjamini Hochberg algorithm. FDR

< 0.05 was considered significant. All analyses were performed in R version 3.6.1.

## Results

### Study design

We collected the RNA-seq data of 504 low-grade-glioma patients separating patients into two groups: younger (age below 50 years) and older patients (age more than 50 years). We further performed differential expression analysis and pathway enrichment analysis to characterize the gene expression differences between the two groups. Finally, we compare metablic and immune gene expression between the younger patients and older patients (**Figure 1**).

**Figure 1.**
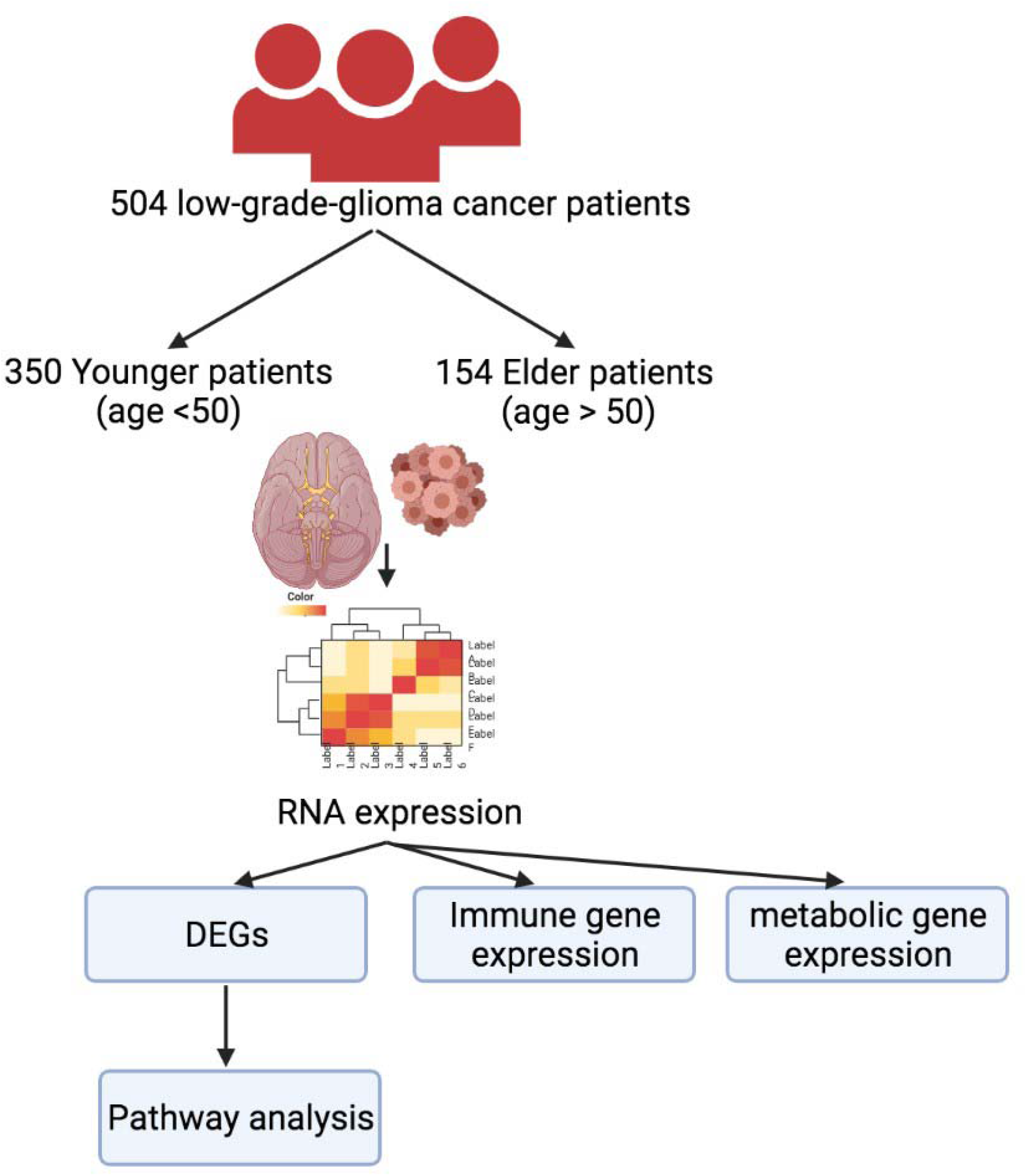
Schema of study design.

### Differentially expressed genes

To characterize the age-related gene-expression alterations, we performed differential expression analysis between younger and older patients’ LGG tissues. We identified 892 up-regulated genes and 1481 down-regulated genes by comparing the expression profiles of 20,282 human coding genes between younger versus older LGG (**Figure 2**). Genes such as ESX1, PRSS35, SFRP2, TPTE22, MKX, IRX2, and PCDH15 were significantly overexpressed (fold change > 1.5) in younger patients, while genes such as ETV4, SPRY4, RIN1, EN1, CHI3L1, and TBX5 were down-regulated (fold change <1.5) in younger patients.

**Figure 2.**
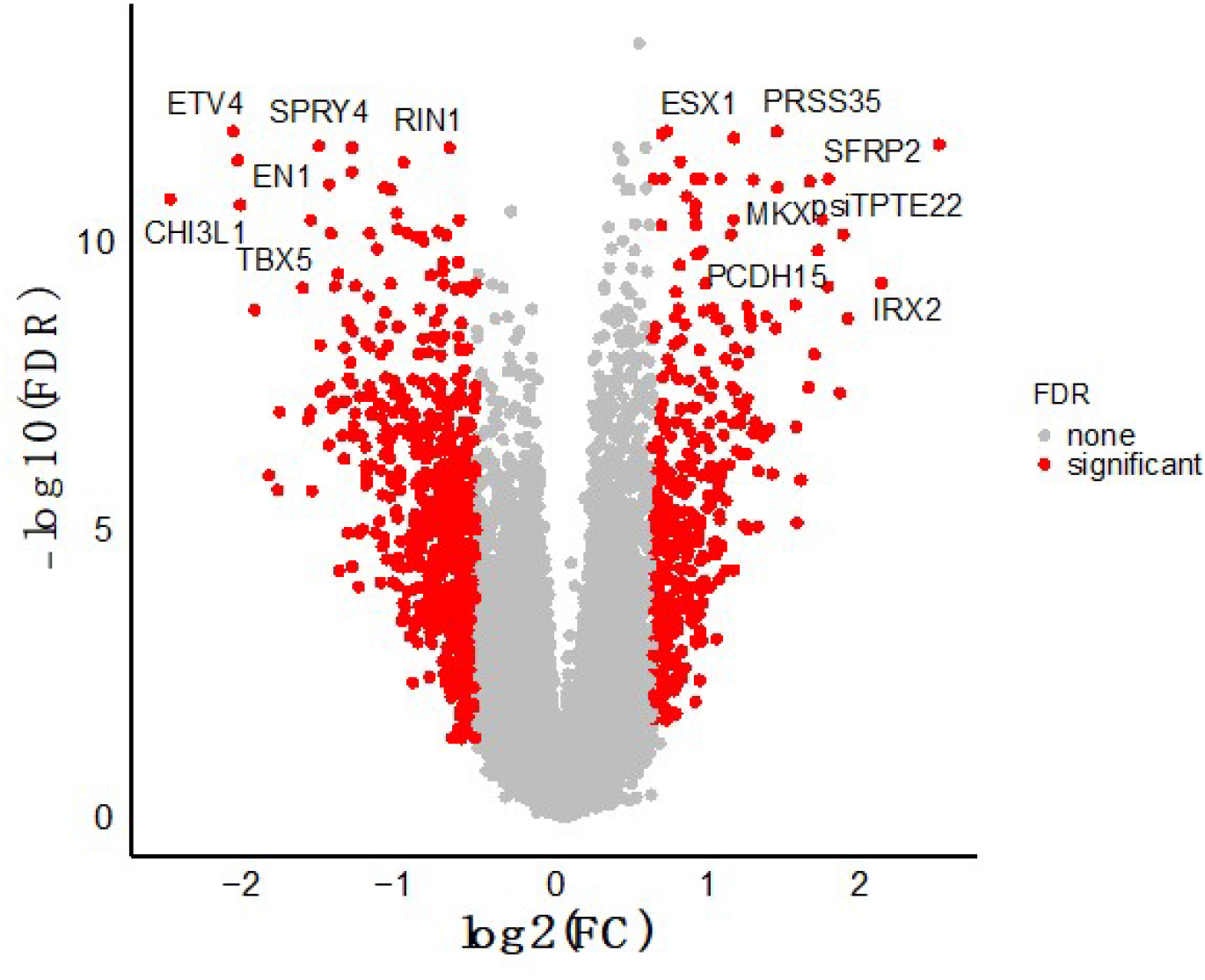
The differentially expressed genes between younger versus older patients in LGG. Volcano plots indicate the fold change (FC) and FDR of differentially expression analysis. P-values were generated by Student’s t-test and corrected by Benjamini Hochberg FDR. Red dots indicated genes meet criteria of |log2 fold change| ≥ 0.585 and FDR < 0.05 (significant), and other genes were marked as grey (none, not significant). Top DEGs with |log2 fold change| > 0.585 and FDR < 1e-05 were labeled with official gene symbols.

### Pathway enrichment analysis

The differentially expressed genes were significantly enriched in 7 KEGG pathways, including protein digestion and absorption, transcriptional misregulation in cancer, ECM-receptor interaction, neuroactive ligand-receptor interaction, amoebiasis, cytokine-cytokine receptor interaction, and complement and coagulation cascades pathways (FDR < 0.05) (**Figure 3**).

**Figure 3.**
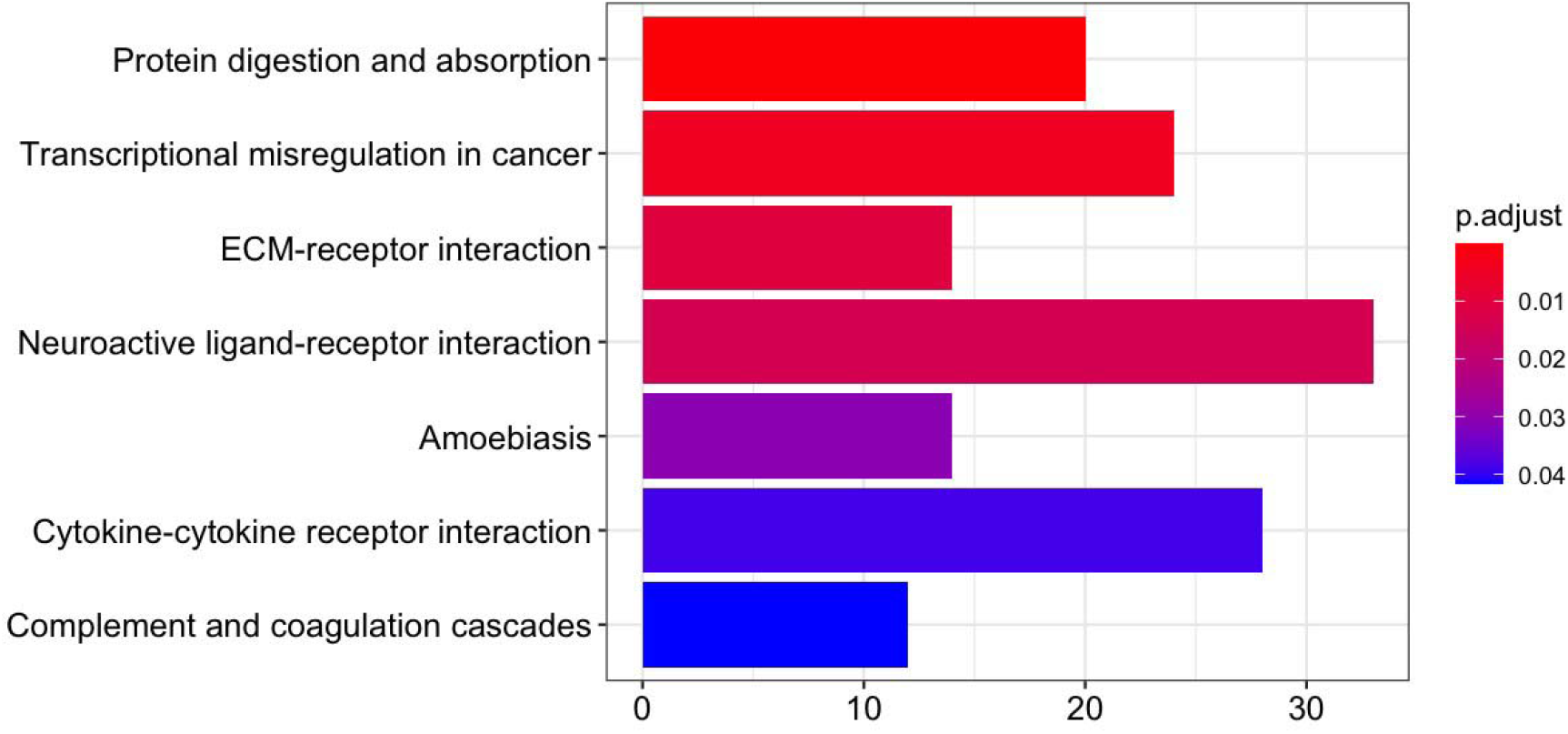
The pathway enrichment analysis of differentially expressed genes. Pathway enrichment p-values were generated by the Fisher’s exact-test and adjusted by Benjamini Hochberg for multiple comparisons. Color indicates the adjusted p value. X-axis indicates the number of differentially expressed genes involved in each pathway. Y-axis indicates the name of enriched KEGG pathways.

### Cancer-related metabolic gene expression in younger versus older LGG patients

We further examined the metabolic gene expression signatures compared between younger and older groups. The gene signatures were defined as the mean of average normalized expression of the member genes and transformed to z-score (Supplementary Table 1). Eleven metabolic signatures were lower expressed in the younger groups compared to the older group, e.g., the nucleotide synthesis, KEAP1 NRF2, and mTOR signatures, while two signatures (autophagy and mTOR) were up-regulated in the younger patients (FDR = 1.21×10-4) (**Table 1**). The mTOR and autophagy genes are upregulated in younger patients while others are down regulated.

**Table 1.** Metabolic signatures associated with age of LGG diagnosis

| Signature | log2 Fold Change | Mean in Young | Mean in Old | P value | FDR |
| --- | --- | --- | --- | --- | --- |
| Nucleotide.Synthesis | -0.58 | -0.18 | 0.40 | 8.36E-09 | 3.51E-07 |
| KEAP1.NRF2.Pathway | -0.44 | -0.14 | 0.31 | 1.34E-06 | 2.81E-05 |
| Autophagy | 0.38 | 0.12 | -0.26 | 1.44E-05 | 1.21E-04 |
| mTOR | 0.38 | 0.12 | -0.27 | 1.21E-05 | 1.21E-04 |
| Tryptophan.Kynurenine.Metabolism | -0.44 | -0.13 | 0.30 | 9.35E-06 | 1.21E-04 |
| Pentose.Phosphate.Pathway | -0.40 | -0.12 | 0.28 | 7.98E-05 | 5.59E-04 |
| Endocytosis | -0.44 | -0.14 | 0.31 | 1.72E-04 | 1.03E-03 |
| DNA.Damage.Repair | -0.34 | -0.11 | 0.24 | 3.80E-04 | 1.99E-03 |
| Cell.Cycle | -0.32 | -0.10 | 0.22 | 8.10E-04 | 3.09E-03 |
| Nucleotide.Salvage | -0.32 | -0.10 | 0.22 | 1.07E-03 | 3.76E-03 |
| Lysosomal.Degradation | -0.36 | -0.11 | 0.25 | 1.19E-03 | 3.84E-03 |
| Amino.Acids.Transporters | -0.29 | -0.09 | 0.20 | 1.64E-03 | 4.93E-03 |
| Reactive.Oxygen.Response | -0.28 | -0.08 | 0.19 | 2.33E-03 | 0.01 |
| Amino.Acids.Synthesis | -0.24 | -0.07 | 0.17 | 0.02 | 0.05 |
| IDH1.2.Activity | -0.24 | -0.07 | 0.16 | 0.02 | 0.05 |
| Fatty.Acids.Synthesis | -0.24 | -0.07 | 0.17 | 0.03 | 0.07 |
| Vitamin...Cofactor.Metabolism | -0.14 | -0.04 | 0.10 | 0.04 | 0.08 |
| PI3K | -0.27 | -0.08 | 0.19 | 0.05 | 0.10 |
| Glycolysis | -0.12 | -0.04 | 0.08 | 0.06 | 0.11 |
| Hypoxia | -0.26 | -0.08 | 0.18 | 0.08 | 0.13 |
| p53.Pathway | -0.12 | -0.04 | 0.08 | 0.07 | 0.13 |
| Myc | -0.18 | -0.06 | 0.13 | 0.09 | 0.16 |
| Antigen.Presentation | -0.20 | -0.06 | 0.14 | 0.10 | 0.16 |
| Cytokine...Chemokine.Signaling | -0.21 | -0.06 | 0.15 | 0.11 | 0.17 |
| TCR...Costimulatory.Signaling | -0.25 | -0.08 | 0.17 | 0.12 | 0.18 |
| Epigenetic.Regulation | 0.19 | 0.06 | -0.13 | 0.15 | 0.22 |
| Mitochondrial.Respiration | -0.12 | -0.04 | 0.08 | 0.22 | 0.31 |
| Transcriptional.Regulation | -0.12 | -0.04 | 0.09 | 0.31 | 0.40 |
| MAPK | 0.06 | 0.02 | -0.04 | 0.35 | 0.44 |
| Arginine.Metabolism | -0.07 | -0.02 | 0.05 | 0.44 | 0.53 |
| Glutamine.Metabolism | 0.09 | 0.03 | -0.06 | 0.44 | 0.53 |
| Fatty.Acids.Oxidation | 0.09 | 0.03 | -0.06 | 0.62 | 0.70 |
| AMPK | 0.00 | 0.00 | 0.00 | 0.64 | 0.71 |
| TLR.Signaling | 0.03 | 0.01 | -0.02 | 0.85 | 0.92 |
| Glucose.Transport | 0.03 | 0.01 | -0.02 | 0.90 | 0.94 |
| NF_Β | -0.04 | -0.01 | 0.03 | 0.94 | 0.96 |

### Cancer-related immune gene expression in younger versus older LGG patients

We finally investigated the MKS proliferation and six immune gene signatures in younger versus older patients including TIS, T Cell, B Cell, Dendritic Cell, and Mast Cell. MKS, TIS, and T Cell signatures are down regulated in young LGG patients while are up regulated in older LGG patients (**Table 2**, FDR<0.02). The results suggest LGG in older patients have higher proliferation activity and immune activity than young patients.

**Table 2.** Immune signatures associated with age of LGG diagnosis

| Signature | log2 Fold Change | Mean in Young | Mean in Old | Pvalue | FDR |
| --- | --- | --- | --- | --- | --- |
| MKS | -0.33 | -0.10 | 0.23 | 6.24E-04 | 2.62E-03 |
| TIS | -0.38 | -0.12 | 0.26 | 5.64E-04 | 2.62E-03 |
| TCell | -0.33 | -0.10 | 0.23 | 0.01 | 0.02 |
| BCell | -0.08 | -0.02 | 0.05 | 0.99 | 0.99 |
| DendriticCell | -0.07 | -0.02 | 0.05 | 0.59 | 0.69 |
| MastCell | -0.07 | -0.02 | 0.05 | 0.23 | 0.31 |

## Discussion

In this study, we characterized the difference in tumor gene expression between younger and older LGG patients. We revealed that DEGs are mostly involved in cancer related pathways. We also identified that older LGG patients have higher metabolic gene expression, immune activity, and proliferation than older patients.

Several DEGs play very important roles in LGG progression according to recent publications. For example, EN1 expression was associated with to LGG prognosis which predicts significantly shorter overall survival time [12]. High expression of CHI3L1 contributed to worse survival [13]. In our analysis, we demonstrated that EN1 and CHI3L1 were highly expressed in older patients which might account for the poor clinical outcome.

The metabolic pathways were up regulated in older patients. Abnormal metabolism is one of the key hallmarks of cancer [14]. Increased consumption of glucose and production of lactate in the presence of oxygen, known as the Warburg effect, has long been appreciated as a metabolic phenotype of cancer [15]. The increasing metabolic activity associated with cancer progression and metastasis [16]. We also demonstrated that older patients have higher proliferation gene expression which is associated with tumor progression and invasion. Our study suggests that the higher metabolic and proliferation gene expression may contribute to the poor clinical prognosis in older LGG patients. Moreover, researches also demonstrated that metabolic, immune, and proliferation gene expression profiling can be used to predict the response to anti-tumor drugs [17, 18]. The higher expression of those signatures in older patients may serve as potential biomarkers for age-related therapeutic response in clinical.

Overall, our study characterizes the gene expression difference of young and old LGG, explaining the molecular mechanism of age-related clinical outcomes which is likely to contribute to its impact on reducing the risk of tumor progression in the older patients.

## Data Availability

All data produced in the present study are available upon reasonable request to the authors

## Authors’ contributions

SBZ and TQ designed the study. All authors participated in the analysis of the data, writing and review of the manuscript.

## Acknowledgments

We would like to give special thanks to Mucheng He, Peiyuan Song, Jinkun Wu, and Zhetian Xue for contributions to calculation, data analysis, and discussion.

